# Scaling diagnostics in times of COVID-19: Colorimetric Loop-mediated Isothermal Amplification (LAMP) assisted by a 3D-printed incubator for cost-effective and scalable detection of SARS-CoV-2

**DOI:** 10.1101/2020.04.09.20058651

**Authors:** Everardo González-González, Itzel Montserrat Lara-Mayorga, Iram Pablo Rodríguez-Sánchez, Felipe Yee-de León, Andrés García-Rubio, Carlos Ezio Garciaméndez-Mijares, Gilberto Emilio Guerra-Alvarez, Germán García-Martínez, Juan Andrés Aguayo-Hernández, Eduardo Márquez-García, Yu Shrike Zhang, Sergio O. Martínez-Chapa, Joaquín Zúñiga, Grissel Trujillo-de Santiago, Mario Moisés Alvarez

## Abstract

By the third week of June 2020, more than 8,500,000 positive cases of COVID-19 and more than 450,000 deaths had been officially reported worldwide. The COVID-19 pandemic arrived in Latin America, India, and Africa—territories in which the mounted infrastructure for diagnosis is greatly underdeveloped. Here, we demonstrate the combined use of a three-dimensional (3D)-printed incubation chamber for commercial Eppendorf PCR tubes, and a colorimetric embodiment of a loop-mediated isothermal amplification (LAMP) reaction scheme for the detection of SARS-CoV-2 nucleic acids. We used this strategy to detect and amplify SARS-CoV-2 genetic sequences using a set of in-house designed initiators that target regions encoding the N protein. We were able to detect and amplify SARS-CoV-2 nucleic acids in the range of 62 to 2 × 10^5^ DNA copies by this straightforward method. Using synthetic SARS-CoV-2 samples and a limited number of RNA extracts from patients, we also demonstrate that colorimetric LAMP is a quantitative method comparable in diagnostic performance to RT-qPCR. We envision that LAMP may greatly enhance the capabilities for COVID-19 testing in situations where RT-qPCR is not feasible or is unavailable. Moreover, the portability, ease of use, and reproducibility of this strategy make it a reliable alternative for deployment of point-of-care SARS-CoV-2 detection efforts during the pandemics.

## Introduction

By the end of the third week of June 2020, more than 8.5 million positive cases of COVID-19 were officially reported across the globe[1]. Even developed countries, such as the USA, England, France, and Germany, are still struggling to mitigate the propagation of SARS-CoV-2 by implementing social distancing and widespread testing. Less developed regions, such as Latin America, India, and Africa, are now experiencing the arrival of COVID-19, but these—territories are woefully lacking in the finances or the mounted infrastructure for diagnosis of this pandemic infection. Rapid and massive testing of thousands of possibly infected subjects has been an important component of the strategy of the countries that are effectively mitigating the spreading of COVID-19 among their populations (i.e., China[2], South Korea [3], and Singapore [4]). By comparison, developing countries with high demographic densities, such as México [5], India [6], or Brazil [7], may not be able to implement a sufficient number of centralized laboratories for rapid large-scale testing for COVID-19.

Many methodologies have been proposed recently to deliver cost-effective diagnosis (i.e., those based on immunoassays [8–11] or specific gene hybridization assisted by CRISPR-Cas systems [12–14]). While immunoassays are an accurate and efficacious tool for assessing the extent of the infection for epidemiological studies [15], their usefulness is limited to the identification of infected subjects during early phases of infection [11,16], a critical period for infectiveness. For instance, experimental evidence collected from a small number of COVID-19 patients (9 subjects) showed that 100% of them produced specific immunoglobulins G (IgGs) for SARS-CoV-2 within two weeks of infection, but only 50% of them did during the first week post infection [17].

Nucleic acid amplification continues to be the gold standard for the detection of viral diseases in the early stages [18–22], and very small viral loads present in symptomatic or asymptomatic patients can be reliably detected using amplification based technics, such as polymerase chain reaction (PCR) [23–25], recombinase polymerase amplification (RPA)[26], and loop-mediated isothermal amplification (LAMP) [27–29].

During the last two pandemic events with influenza A/H1N1/2009 and COVID-19, the Centers for Disease Control and Prevention (CDC) and the World Health Organization (WHO) recommended real-time quantitative PCR (RT-qPCR) methods as the gold standard for official detection of positive cases[16,30]. However, the reliance on RT-qPCR often leads to dependence on centralized laboratory facilities for testing [16,30–33]. To resolve this drawback, isothermal amplification reaction schemes (i.e., LAMP and RPA) have been proposed as alternatives to PCR-based methods and devices for point-of-care settings [32,34,35]. The urgency of using reliable molecular-based point of care (POC) methods for massive diagnostic during epidemiological emergencies has become even more evident during the current COVID-19 pandemics [30,36,37].

In these times of COVID-19 [38], scientists and philanthropists around the globe have worked expeditiously on the development of rapid and portable diagnostics for SARS-CoV-2. Several reports have demonstrated the use of colorimetric LAMP-based methods for diagnosis of pandemic COVID-19 [39–44]. Some of these reports (currently available as preprints) use phenol red, a well-known pH indicator, to assist in the visual discrimination between positive and negative samples [39,40,45].

In this study, we demonstrate the use of a simple embodiment of a colorimetric LAMP protocol for the detection and amplification of synthetic samples and actual RNA samples from patients of SARS-CoV-2, the causal viral agent of COVID-D. In this LAMP-based strategy, also assisted by the use of phenol red, sample incubation is greatly facilitated by the use of a three-dimensional (3D)-printed incubator connected to a conventional water circulator, while discrimination between positive and negative samples is achieved by visual inspection. We quantitatively analyze differences in color between positive and negative samples using color decomposition and analysis in the color CIELab space[46]. Moreover, we compare the sensibility of this LAMP colorimetric method versus PCR protocols. This simple strategy is potentially adequate for the fast deployment of diagnostic efforts in the context of COVID-19 pandemics.

### Rationale

We have developed a simple diagnostic method for the detection of SARS-CoV-2, the causal agent of COVID-19. The rationale underlying this strategy is centered on achieving the simplest possible integration of easily available reagents, materials, and fabrication techniques to facilitate fast and massive implementation during the current COVID-19 pandemics in low- or middle-income regions. This method is based on the amplification of the genetic material of SARS-CoV-2 using LAMP. The amplification is conducted using a commercial reaction mix in commercial and widely available 200 µL Eppendorf PCR tubes. Moreover, we have designed and fabricated a simple 3D-printed chamber (Figure 1) for incubation of the Eppendorf tubes and to enable LAMP at high temperatures (50–65 °C) and extended times (up to 1 h). We show that this incubation chamber, when connected to a conventional water recirculator, enables the successful amplification of positive samples (i.e., samples containing SARS-CoV-2 nucleic acids).

**Figure 1.**
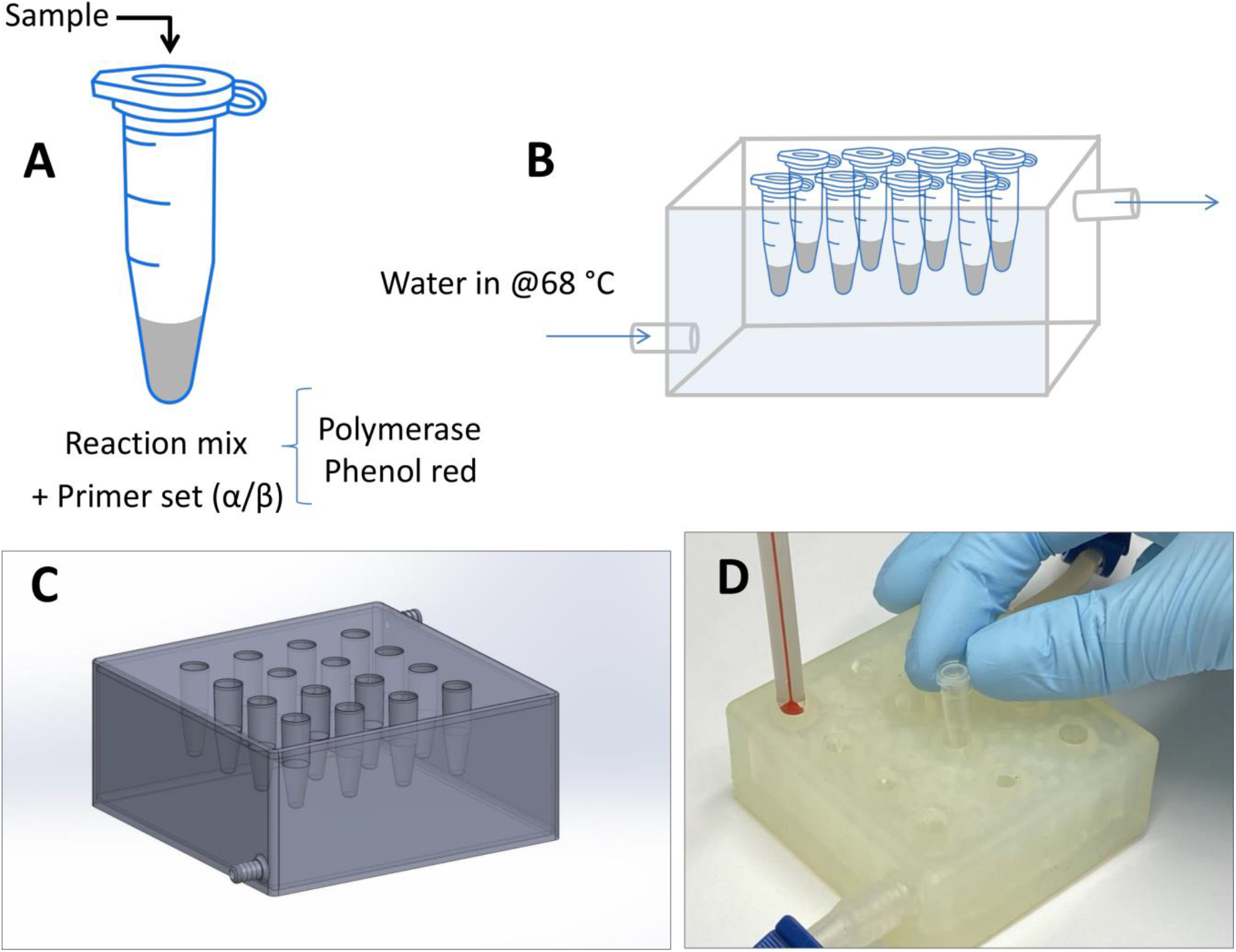
Experimental setup. (A) Commercial 200 microliter Eppendorf PCR tubes, and (B) a 3D-printed incubator was used in amplification experiments of samples containing the synthetic SARS-CoV-2 nucleic acid material. (C) 3D CAD model of the LAMP reaction incubator. (D) Actual image of the Eppendorf tube incubator connected to a conventional water circulator.

This incubation chamber is one of the key elements that enable rapid and widespread implementation of this diagnostic method at low cost. This 3D-printed incubator can be rapidly printed using standard SLA printers widely available in markets worldwide.

Standard 3D-printing resins can be used. The availability of the original AutoCAD files (included here as supplemental material) enables fast modification/optimization of the design for accommodation of a larger number of samples or larger or smaller tubes, adaptation to any available hoses (tubing), and possible incorporation of an on-line color-reading system.

Indeed, all this is consistent with the main rationale of our proposed diagnostic strategy for pandemic COVID-19: To enabling a fast and feasible response using widespread, distributed, and scalable diagnostics fabricated with widely available resources.

In the following section, we briefly discuss the mechanisms of amplification and visual discrimination between positive and negative samples.

### Colorimetric LAMP amplification

The presence of phenol red within the LAMP reaction mix allows for naked-eye discrimination between positive and negative samples (Figure 2). The reaction mix is coupled with the pH color transition of phenol red, a widely used pH indicator, which shifts in color from red to yellow at pH 6.8. During LAMP amplification, the pH of the reaction mix continuously evolves from neutrality to acidic values as protons are produced [27,47]. The mechanism of production of hydrogen ions (H^+^) during amplification in weakly buffered solutions has been described [47]. DNA polymerases incorporate a deoxynucleoside triphosphate into the nascent DNA chain. During this chemical event, a pyrophosphate moiety and a hydrogen ion are released as byproducts (Figure 2A). This release of hydrogen ions is quantitative, according to the reaction scheme illustrated in Figure 2. The caudal of H^+^ is high, since it is quantitatively proportional to the number of newly integrated dNTPs. In fact, the quantitative production of H^+^ is the basis of previously reported detection methods, such as the semiconductor sequencing technology operating in Ion Torrent sequencers[48]. In the initially neutral and weakly buffered reaction mixes, the production of H^+^ during LAMP amplification progressively and rapidly shifts the pH across the threshold of phenol red (Figure 2B).

**Figure 2.**
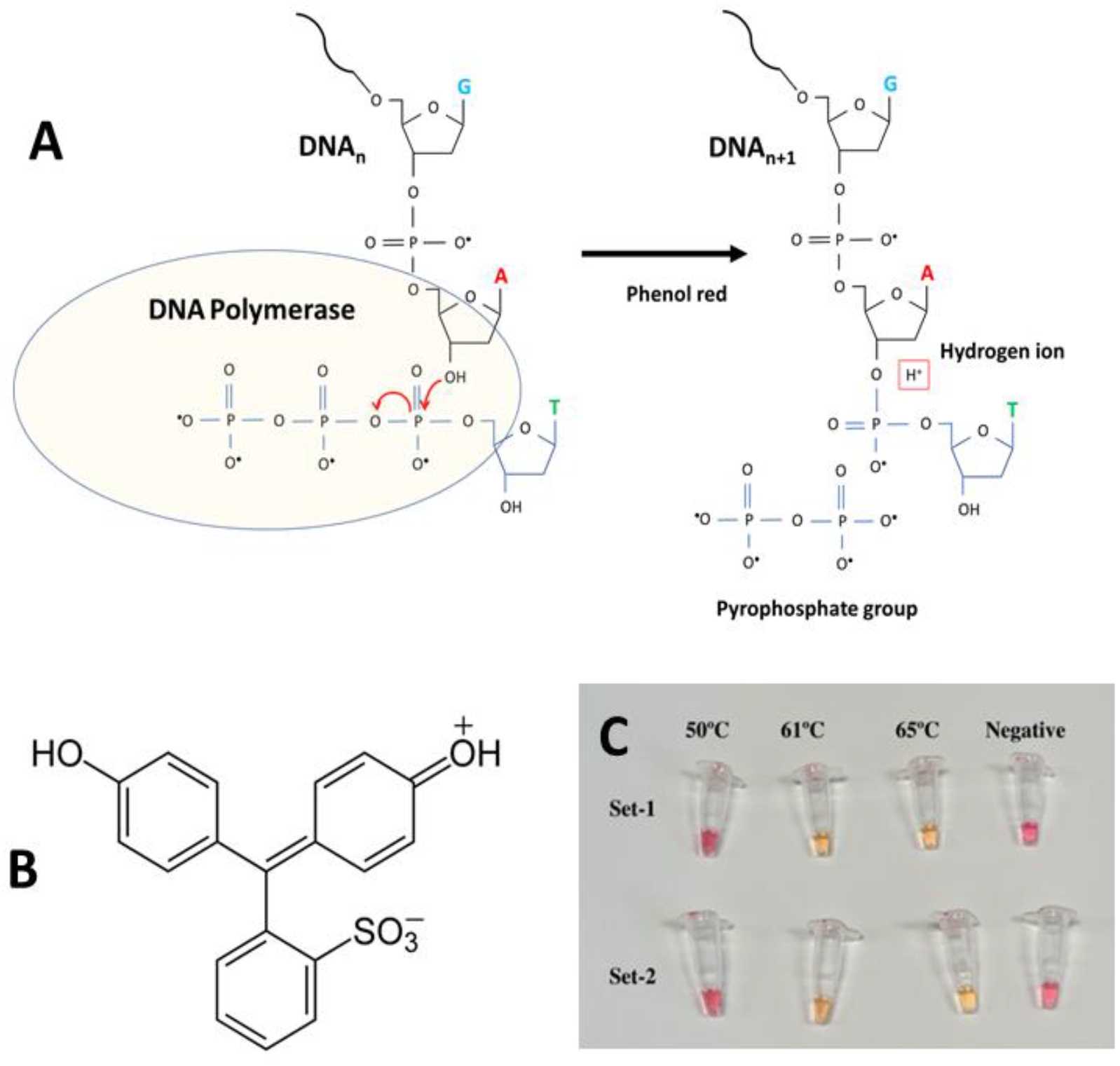
Initiators and pH indicator for SARS-Co2 detection using a colorimetric LAMP method. (A) The LAMP reaction scheme. (B) Chemical structure of phenol red. (C) Two different sets of LAMP primers were used for successfully targeting a gene sequence encoding the SARS-Co2 N protein. Successful targeting and amplification are clearly evident to the naked eye: positive samples shift from red to yellow.

Moreover, the pH shift is clearly evident to the naked eye, thereby freeing the user from reliance on spectrophotometric instruments and facilitating simple implementation during emergencies (Figure 2C). Images in Figure 2C show representative colors of the amplification reaction mixes contained in Eppendorf PCR tubes after incubation for 30 min. Three different incubation temperatures were tested (50, 60, and 65 °C) and two different sets of LAMP-primers (α and β) were used (Table 1).

**Table 1.**
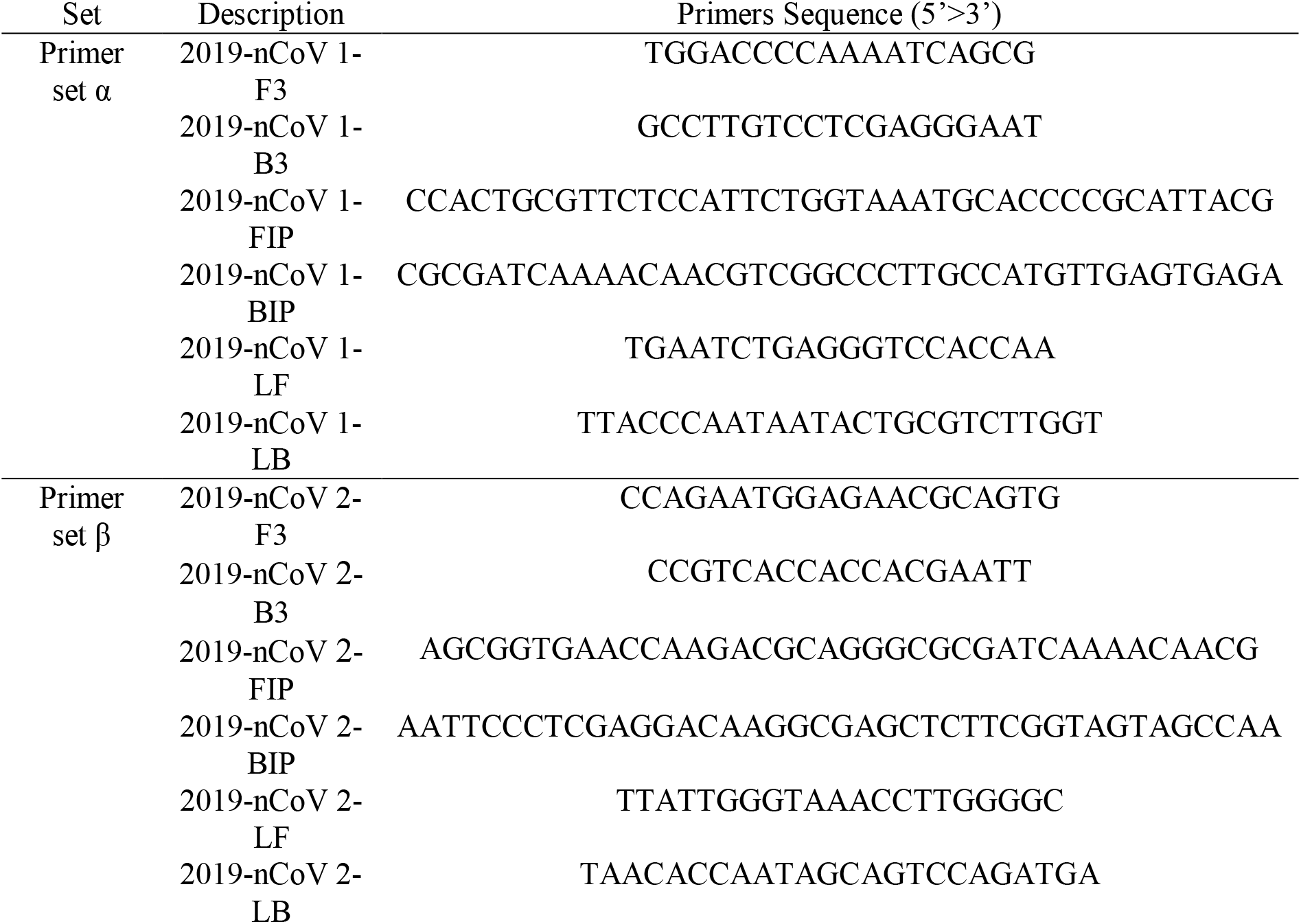
Primer sequences used in LAMP amplification experiments. Two different sets of primers were used, directed at the RNA sequence encoding the N sequence of the SARS-CoV-2.

Both sets of primers performed equivalently, at least based on visual inspection, in the three temperature conditions tested. Discrimination between positive and negative controls is possible using only the naked eye to discern the reaction products from amplifications conducted at 60 and 65 °C. No or negligible amplification was evident at 50 °C or in the control group.

Furthermore, we were able to successfully discriminate between positive and negative samples using LAMP reaction mix already added with primers and kept at 20 °C or 4 °C for 24, 48, 72, and 96 h (Figure S5). The stability of the reaction, the isothermal nature of the amplification process, and its independence from specialized equipment greatly simplifies the logistic of implementation of this diagnostic method outside centralized labs.

### Analysis of sensitivity

We conducted a series of experiments to assess the sensitivity of the LAMP reactions in the 3D-printed incubation chamber using the two sets of primers (α and β; Table 1). The amplification proceeds with sufficient quality to also allow proper visualization of the amplification products in electrophoresis gels, even at low nucleic acid concentrations. We observed that amplification proceeded successfully in a wide range of viral loads, from 625 to 5 × 10^5^ copies in experiments using synthetic SARS-CoV-2 nucleic acid material (Figure 3A). We clearly observed amplification in samples containing as few as 625 viral copies after incubation times of 50 min at 65 °C. If we put this range into a proper clinical context, the actual viral load of COVID-19 in nasal swabs from patients has been estimated to fall within the range of 10^5^ to 10^6^ viral copies per mL [49]. Discrimination between positive and negative samples (controls) can be clearly established by the naked eye in all reactions incubated for 50 min, regardless of the number of viral copies present. In addition, we did not observe any non-specific amplification in negative samples (i.e., containing synthetic genetic material form EBOV) incubated for 50 min at 65 °C. Indeed, the identification and amplification of SARS-CoV-2 synthetic material is feasible in samples that contained ∼62.5 viral copies using this LAMP strategy (Figure S3) and incubation times of 50–60 min.

**Figure 3.**
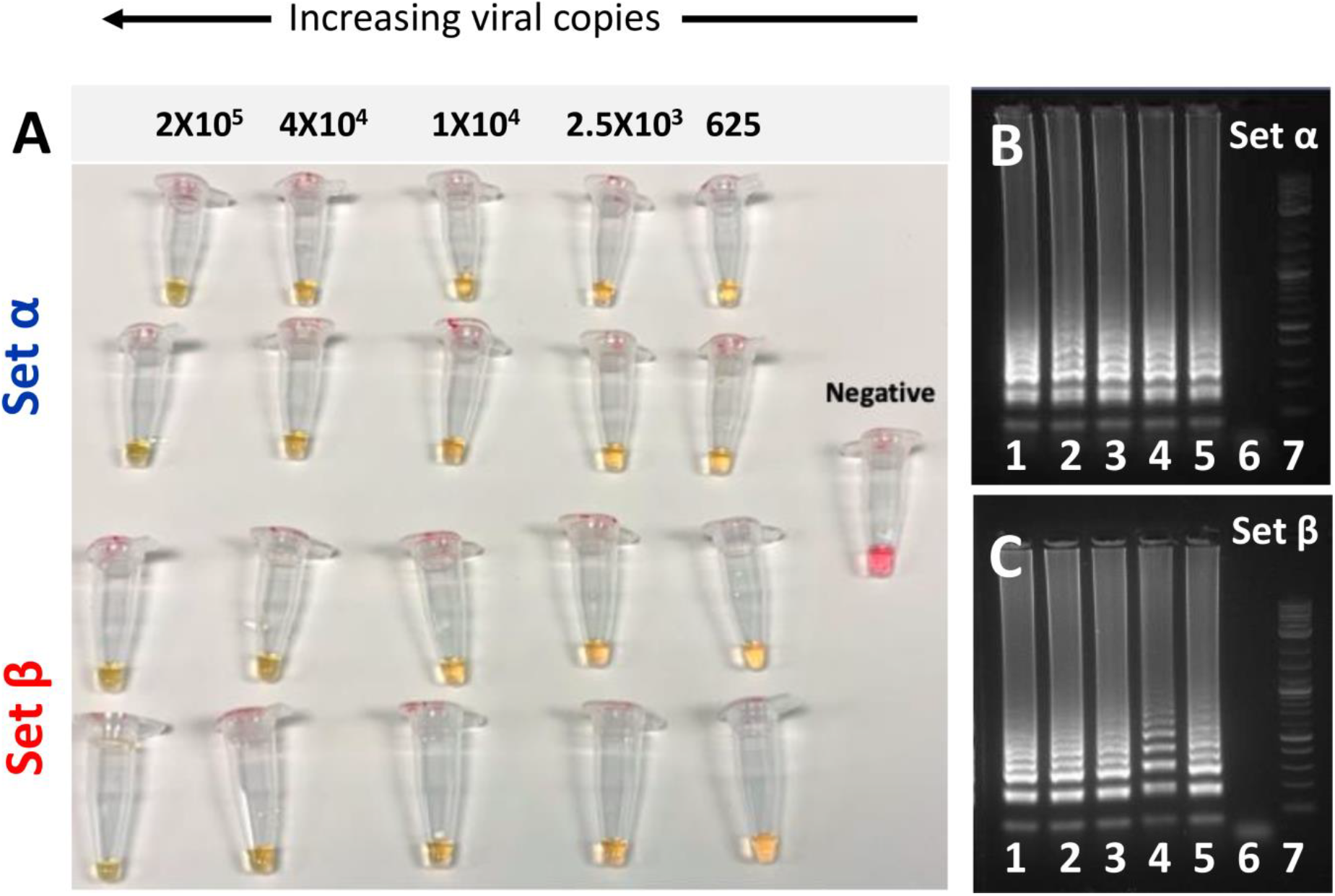
Two different sets of LAMP-primers were used for successfully targeting of a gene sequence encoding the SARS-Co2 N protein. (A) LAMP primer sets α and β both enable the amplification of synthetic samples of SARS-CoV-2 nucleic acids in a wide range of template concentrations, from 625 to 2.0 × 10^5^ DNA copies of SARS-CoV-2 when incubated for 50 minutes at a temperature range from 60 to 65 °C. (B, C) Agarose gel electrophoresis of DNA amplification products generated by targeting two different regions of the sequence coding for SARS-Co2 N protein. Two different primer sets were used: (B) primer set α, and (C) primer set β. The initial template amount was gradually decreased from left to right: 2.0 × 10^5^ DNA copies (lane 1), 4.0 × 10^4^ copies, (lane 2), 1.0 × 10^4^ copies (lane 3), 2.5 × 10^3^ copies (lane 4), 625 copies (lane 5), negative control (lane 6), and molecular weight ladder (lane 7). Panel B and C corresponds to portions of the full-length gels presented in supplemental figures S8A and S8B, respectively.

We corroborated the amplification by visualizing LAMP products with gel electrophoresis for the different viral loads tested. Figures 3B, C show agarose gels of the amplification products of each one of the LAMP experiments, where two different sets of primers (α and β) were used to amplify the same range of concentrations of template (from 625 to 2 × 10^5^ synthetic viral copies). We were able to generate a visible array of bands of amplification products, a typical signature of LAMP, for both LAMP primer sets and across the whole range of synthetic viral loads. Indeed, both primer sets rendered similar amplification profiles.

In summary, using the primers and methods described here, we were able to consistently detect the presence of SARS-CoV-2 synthetic nucleic acids. We have used a simple 3D-printed incubator, connected to a water circulator, to conduct LAMP. We show that, after only 30 min of incubation, samples containing a viral load in the range of 10^4^ to 10^5^ copies could be clearly discriminated from negative samples by visual inspection with the naked eye (Figure 2C). Samples with a lower viral load were clearly discriminated when the LAMP reaction was incubated for 50 min. Incubation periods of up to 1 h at 68 °C did not induced false positives and were able to amplify as few as ∼62 copies of SARS-CoV-2 synthetic genetic material. These results are consistent with those of other reports in which colorimetric LAMP, assisted by phenol red, has been used to amplify SARS-COV-2 genetic material [39,40]. We observe 0 false positive cases in experiments where synthetic samples containing EBOV genetic material were incubated at 65 °C for 1 h.

In the current context of the COVID-19 pandemics, the importance of communicating this result does not reside in its novelty but in its practicality. Some cost considerations follow. While the market value of a traditional RT-qPCR apparatus (the current gold standard for COVID-19 diagnostics) is in the range of 10,000 to 40,000 USD, a 3D-printed incubator, such as the one described here (Figure S1,S2; Supplementary file S1), could be fabricated for under 200 USD at any 3D printer shop. This difference is significant, especially during an epidemic or pandemic crisis when rational investment of resources is critical. While the quantitative capabilities of testing using an RT-qPCR platform are undisputable, the capacity of many countries to rapidly, effectively, and massively establish diagnostic centers based on RT-qPCR is questionable. The current pandemic scenarios experienced in the USA, Italy, France, and Spain, among others, have crudely demonstrated that centralized labs are not an ideal solution during emergencies. Portable diagnostic systems may provide a vital flexibility and speed of response that RT-qPCR platforms cannot deliver.

### Feasibility of real-time quantification

Here, we further illustrate the deterministic and quantitative dependence between the concentration of the amplification product and the color signal produced during this colorimetric LAMP reaction. For this purpose, we simulated real-time amplification experiments by conducting a series of amplification reactions using initial amounts of 625, 1 × 10^4^, and 2 × 10^5^ copies of synthetic SARS-CoV-2 genetic material in our 3D-printed incubator.

We extracted samples from the incubator after 0, 10, 20, 30, 40, and 50 min of incubation at 65 °C. The color of these samples was documented as images captured using a smart phone (iPhone 7) against a white background (Figure 4A). The images were analyzed using the free access application Color Companion^®^ for the iPhone or iPad. Briefly, color images were decomposed into their CIELab space components. In the CIELab color space, each color can be represented as a point in a 3D-space, defined by the values **L, a**, and **b** [46]. In this coordinate system, **L** is the luminosity (which ranges from 0 to +100), **a** is the blue-yellow axis (which ranges from −50 to 50), and **b** is the green-red axis (which ranges from - 50 to 50) (Figure S4).

**Figure 4.**
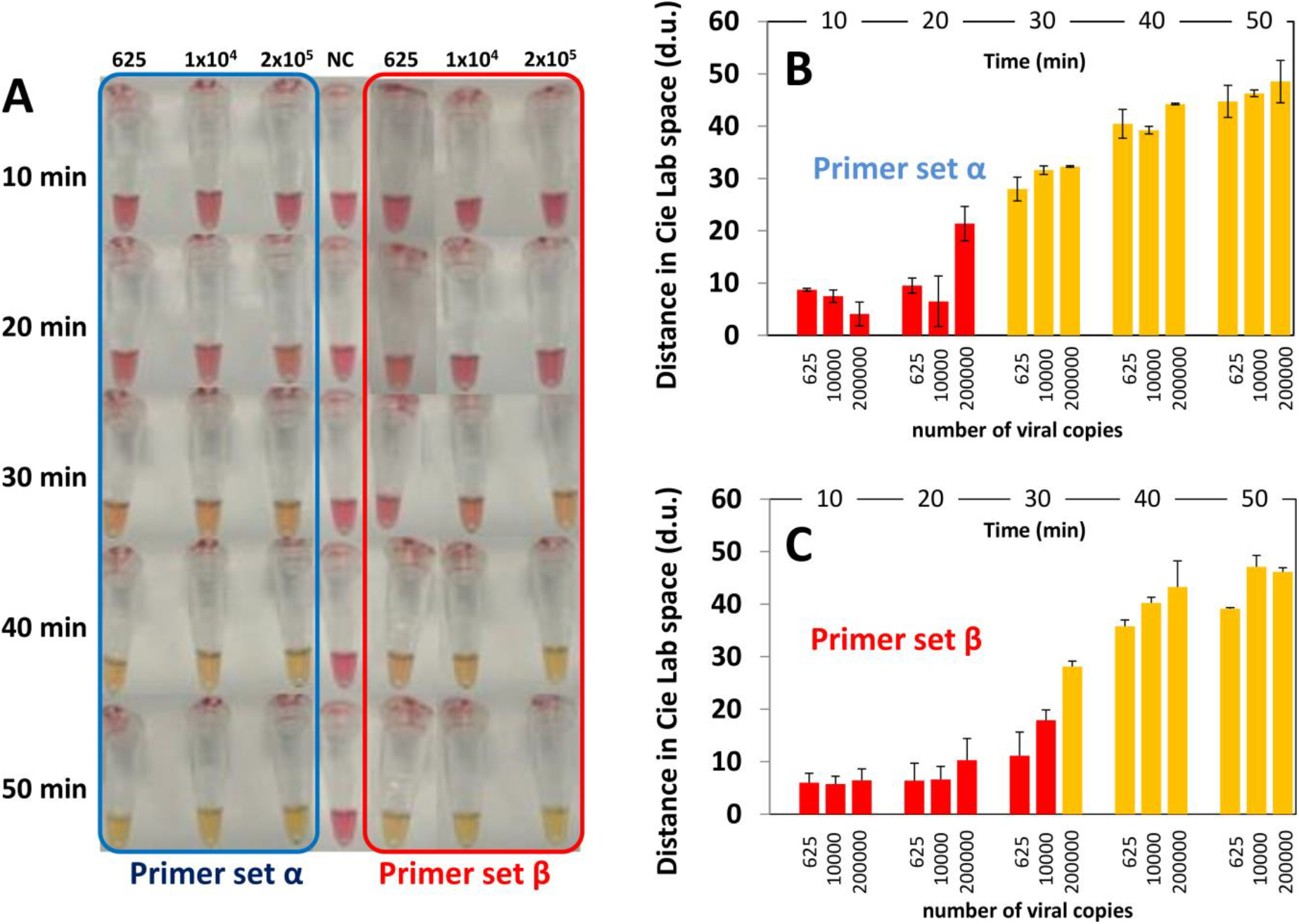
Evaluation of the sensitivity of the combined use of a colorimetric LAMP method assisted by the use of phenol red. (A) Sensitivity trials using different concentrations of the template (positive control) and two different primers sets: α (indicated in blue) and β (indicated in red). Photographs of the Eppendorf PCR tubes containing positive samples and negative controls were acquired using a smartphone. (C, D) Distance in the color CIELab space between negative controls (red) and samples containing different concentrations of SARS-CoV-2 nucleic acid material (i.e., 625, 10000, and 200000 synthetic copies) analyzed after different times of incubation (i.e., 10, 20, 30, 40, and 50 minutes) at 65 °C. The analysis of color distances is presented for amplifications conducted using primer set (B) α and (C) β.

The difference between two colors can be quantitatively represented as the distance between the two points that those colors represent in the CIELab coordinate system. For the colorimetric LAMP reaction mixture used in our experiments, the spectrum of possible colors evolves from red (for negative controls and negative samples) to yellow (for positive samples). Conveniently, the full range of colors for samples and controls can be represented in the red and yellow quadrant defined by L [0,100], a [0,50], and b [0,50]. For instance, the difference between the color of a sample (at any time of the reaction) and the color of the negative control (red; L=53.72 ± 0.581, a=38.86 ± 2.916, and b=11.86 ± 0.961) can be calculated in the CIELab space. We determined the distance in the CIELab space between the color of samples taken at different incubation times that contained SARS-CoV-2 genetic material and negative controls (Figure 4B, C). We repeated this calculation for each of the LAMP primer sets that we used, namely primer set α (Figure 4B) and β (Figure 4C). These results suggest that the color difference between the samples and negative controls is quantifiable. Therefore, color analysis may be implemented to assist the discrimination between positives and negatives. Furthermore, imaging and color analysis techniques may be implemented in this simple colorimetric LAMP diagnostic strategy to render a real-time quantitative Lamp (RT-qLAMP).

Alternatively, the progression of the amplification at different times can be monitored by adding an intercalating DNA agent (i.e., EvaGreen Dye), and measuring fluorescence on time (Figure S5).

Note that the variance coefficients for the control are 1.08, 7.50, and 8.10% for L, a, and b, respectively. These small values suggest robustness and reproducibility in the location of the coordinates of the control point (reference point). Similarly, the variation in color between negative controls and positive samples incubated for 50 min was reproducible and robust (average of 46.60 +/-4.02 d.u.; variance coefficient of 8.62%).

Interestingly, we observed significant differences in the performance of the two LAMP primer sets used in the experiments reported here (Figures 4B and 5). Our results suggest that primer set α enabled faster amplification in samples with fewer viral copies. Consistently, this primer set yielded positive discrimination in samples with 625 viral copies in 30 min (Figure 4B). The use of primer set β enabled similar differences in color, measured as distances in the CIELab 3D-space, in 40 min (Figure 4C).

**Figure 5.**
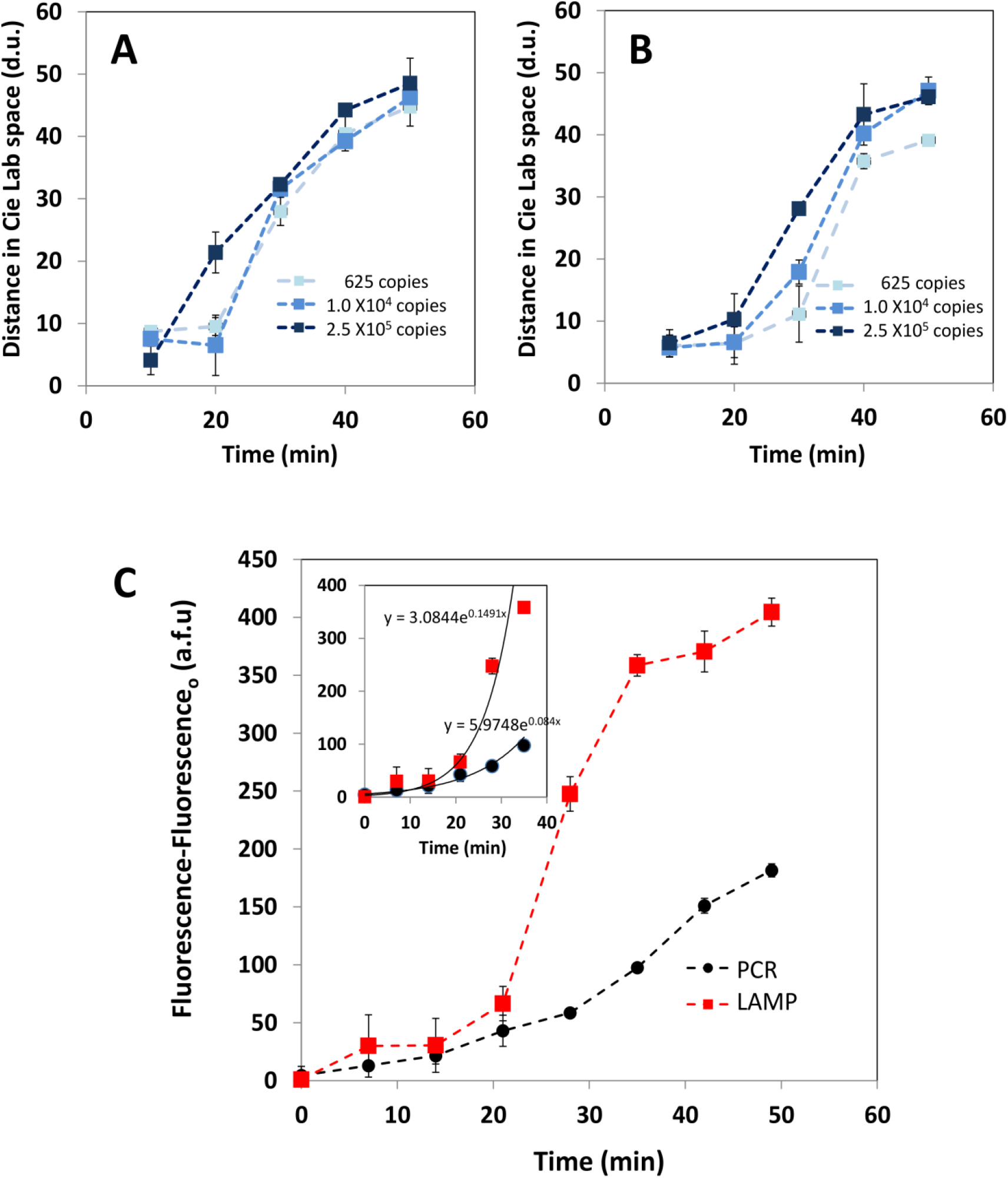
Time progression of the distance in color with respect to negative controls (red color) in the CIELab space for positive SARS-CoV-2 samples containing 625 (light blue, ▪), 1 × 10^4^ (medium blue, ▪), and 2.5 × 10^6^ (dark blue, ▪) copies of synthetic of SARS-CoV-2 nucleic acids. Results obtained in experiments using (A) primer set α, and (B) primer set β. (C) Comparison between the performance of PCR and LAMP in a simulated real-time experiment. Progression of the fluorescence signal, as measured in a plate reader, in PCR (black circles) and LAMP (red squares) experiments. The inset shows a zoom at the exponential stage of the amplification process.

These findings suggest that primer set α should be preferred for final-point implementations of this colorimetric LAMP method. Interestingly, primer set β may better serve the purpose of a real-time implementation. While primer set α produced similar trajectories of evolution of color in samples that contained 1.0 × 10^4^ and 2.0 × 10^5^ viral copies (Figure 5A), primer set β was better at discriminating between amplifications produced from different initial viral loads (Figure 5B).

### Comparison of LAMP versus PCR

LAMP has been regarded before as a more efficient amplification reaction than PCR, since more DNA is produced per unit of time due to the use of a higher number of primers[50] (in this case 6 versus 2). To finalize our analysis, we simulated some real-time amplification experiments to compare the performance of LAMP and PCR in similar conditions (Figure 6A). To that end, we conducted amplification reactions using initial amounts of 4 × 10^4^ copies of synthetic SARS-CoV2 in a commercial miniPCR cycler[24,51] (using primer set N1) and in our LAMP 3D-printed incubator (using primer set β). We added the intercalating agent, EvaGreen^®^ Dye, to the reaction mix at the initial time and extracted samples after 0, 7, 14, 21, 28, 35, 42, and 51 minutes. These samples form PCR and LAMP experiments were dispensed in 96-weel plates. The fluorescence from these samples was then measured in a commercial plate reader[24] (Figure 5C). We observed an exponential increase in fluorescence as more LAMP or PCR cycles were performed, which highlights the quantitative nature of the intercalating reaction. The LAMP reaction produces significantly higher fluorescence signals that the PCR reaction throughout the entire reaction time. The difference between the fluorescence emissions of both amplifications is more evident after the first 20 minutes of amplification. These results also suggest that using a commercial plate reader to determine the extent of advance of LAMP amplifications is a practical and reliable alternative to the use of colorimetric evaluations. Moreover, fluorescence reading of LAMP products may lead to precise quantification of SARS-CoV-2 viral loads.

**Figure 6.**
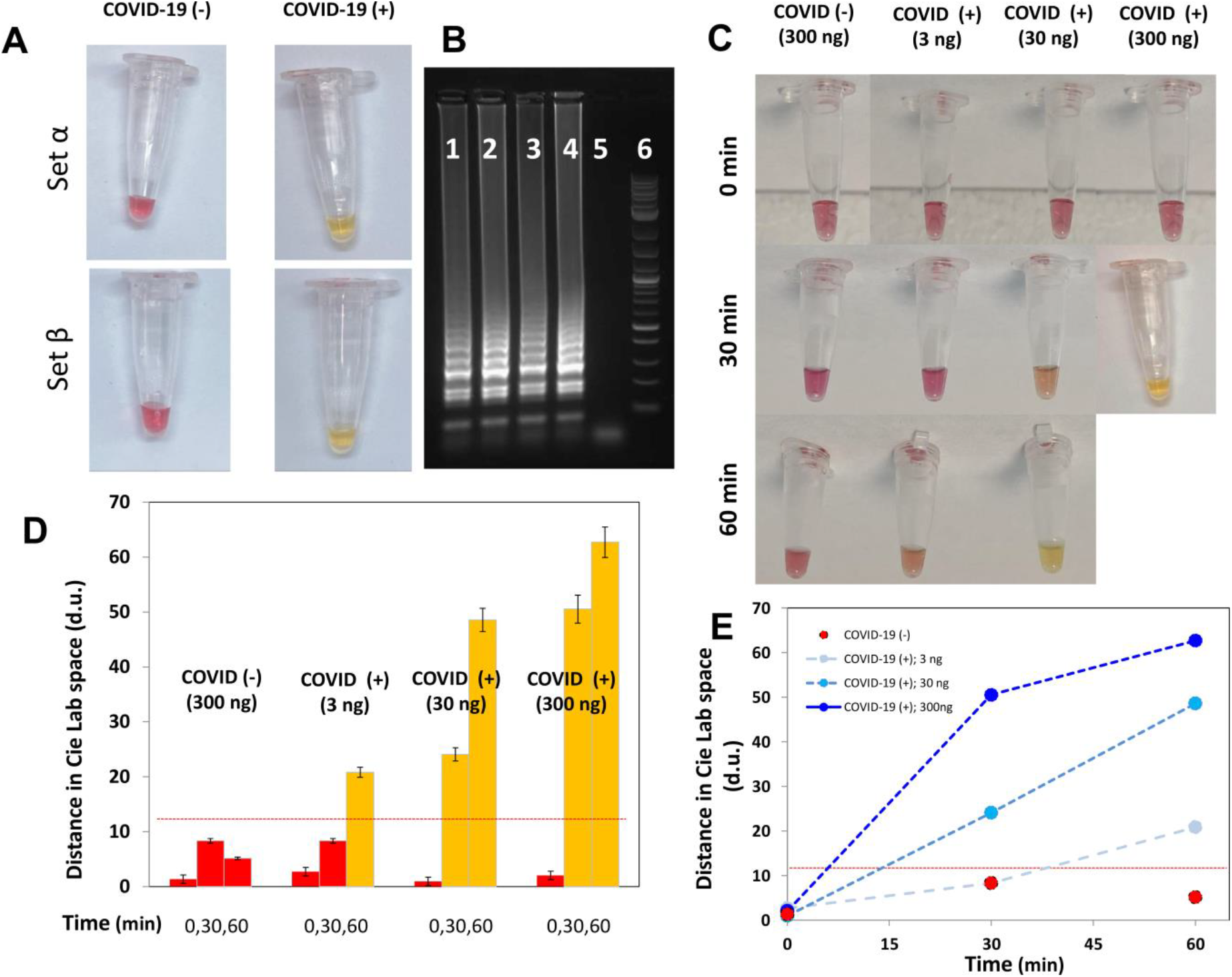
Progression of color changes during amplification in actual RNA extracts from patients. (A) RNA extracts from COVID-19(+) and COVID-19(-) samples, amplified by colorimetric LAMP, can be easily discriminated by visual inspection. (B) LAMP amplification products from RNA (lane 1 and 2) and RNA extracts (lane 3 and 4) from a COVID (+) patient, and a COVID (-) volunteer (lane 5), as revealed by gel electrophoresis experiments. A molecular weight ladder is shown in lane 6. Panel B corresponds to a portion of the full-length gel presented in Supplemental figures (Figure S9). (C) Time progression of color changes in LAMP reaction mixes containing 300 ng of RNA extract from a COVID(-) volunteer (as diagnosed by RT-qPCR), and 3, 30, and 300 ng of RNA extract from a COVID(+) patient (as diagnosed by RT-qPCR). (D) Distance in color with respect to negative controls (red color) in the CIELab space for RNA extracts from a COVID(-) volunteer (as diagnosed by RT-qPCR) containing 300 ng of nucleic acids, and a COVID(+) patient (as diagnosed by RT-qPCR) containing 3, 30, and 35 (dark blue, ▪) ng of nucleic acids. Readings at 0, 30, and 60 minutes are shown. A suggested positive–negative threshold value is indicated with a red line. (E) Time progression of the distance in color with respect to negative controls (red color) in the CIELab space for RNA extracts from a COVID(-) volunteer (as diagnosed by RT-qPCR) containing 300 ng of nucleic acids (red, ▪), and from a COVID(+) patient (as diagnosed by RT-qPCR) containing 3 (light blue, ▪), 30 (medium blue, ▪), and 35 (dark blue, ▪) ng of nucleic acids. A suggested positive–negative threshold value is indicated with a red line.

We also compared the performance of RT-qPCR and colorimetric LAMP using actual RNA extracts isolated from human volunteers. For this purpose, first we used colorimetric LAMP for diagnosis of one RNA sample confirmed as positive for COVID-19 and one confirmed as negative according to RT-qPCR results. RNA extracts from the COVID-19 (+) patient were clearly discriminated from the COVID-19 (-) patient extracts by our colorimetric LAMP amplifications (Figure 6A).

Similar results were obtained regardless of the LAMP primer set used (i.e., α and β). We corroborated our LAMP amplification results using standard gel electrophoresis (Figure 6B). In addition, samples were serially diluted to challenge the sensitivity of colorimetric LAMP. We were able to discriminate between positive and negative samples in the entire concentration range tested (300 ng of total RNA, as determined by nanoDrop assays). The color shift (red to yellow) was clearly perceived after 30 minutes of amplification in samples containing 300 ng of total RNA from COVID (+) patients. Samples containing 30 and 3 ng of total RNA required longer times (Figure 6C).

Positive samples exhibited a shift in color after 60 minutes of amplification, while negative samples remained unchanged. We quantified the change in color in positive and negative samples using color image analysis and by calculating color distances in the CieLab color space (Figure 6D,E).

Our experiments show that the distance in color between positive and negative RNA samples from human volunteers is proportional to the number of viral copies. These results suggest that the change in color can be quantitatively related to the viral load of SARS-CoV-2 in actual RNA extracts, similarly to synthetic samples.

In a final set of experiments, we blind tested a set of 8 extracts of human RNA from nasopharyngeal samples corresponding to 2 patients that were diagnosed as COVID-19 (-) and 6 patients diagnosed as COVID-19 (+) by RT-qPCR. We adjust the RNA content in all samples to 300 ng/µL of RNA and then diluted them to derive samples containing 30 ng/µL. All samples, undiluted and diluted, were added with the LAMP reactive mix and incubated at 65 ° C by 50 minutes. All samples exhibit a red color before incubation (Figure 7A), and only positive samples shifted to yellow during incubation (Figure 7B). We confirmed results by gel electrophoresis of the amplification products. Only positive samples exhibited the characteristic DNA profile associated with LAMP products (Figure 7C,D). Colorimetric LAMP was also able to discriminate positive samples correctly even in diluted extracts containing one order of magnitude less RNA than original extracts. COVID-19 positive RNA samples, original or diluted, showed similar values of distance in color with respect to negative samples, although standard deviations were higher in samples that contained 30 ng/µL than in samples that contained 30 ng/µL (Figure 7E).

**Figure 7.**
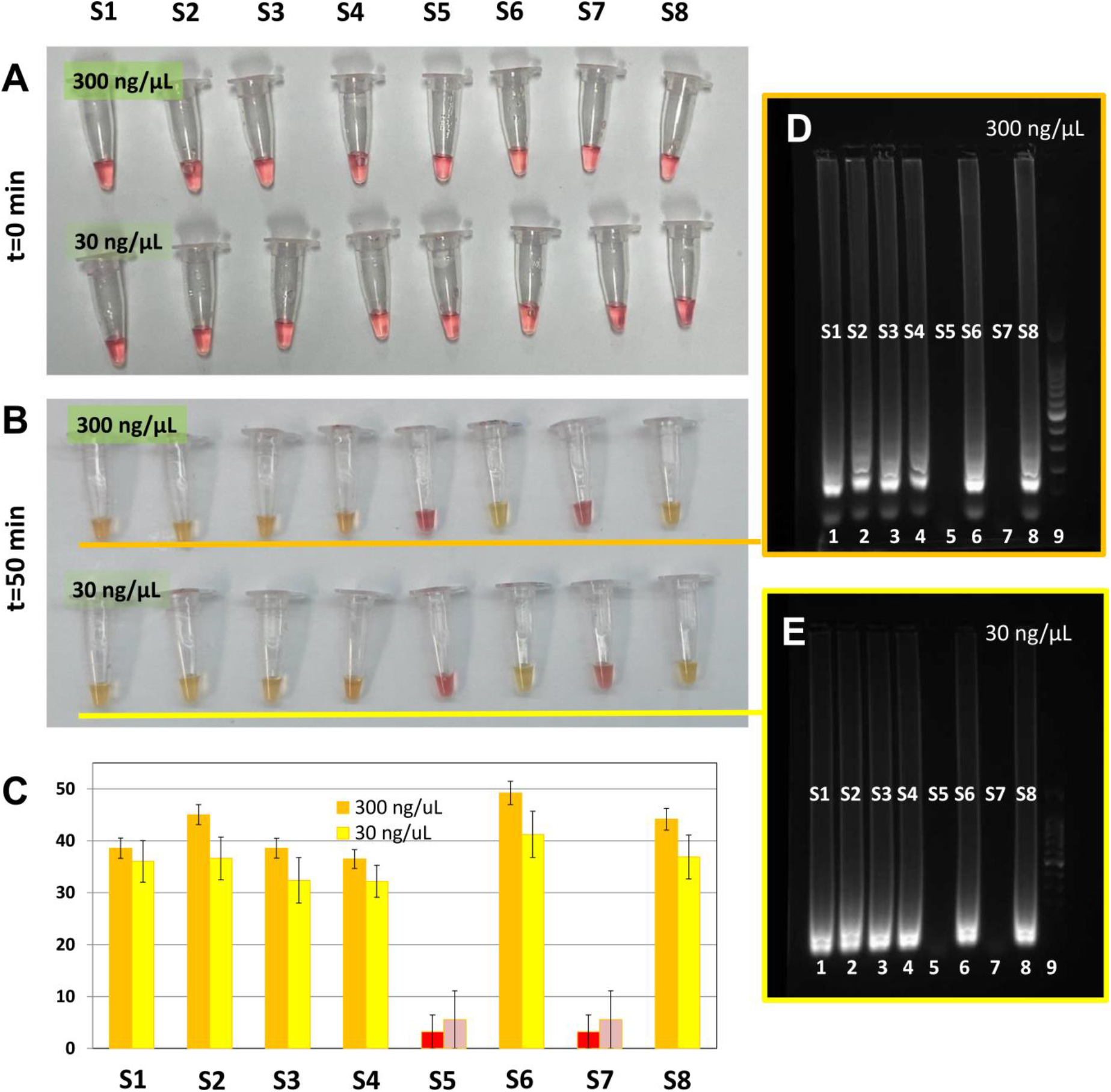
Discrimination of actual RNA extracts from COVID-19 positive and negative samples. Color of RNA extracts from 6 COVID-19(+) and 2 COVID-19(-) samples, at two different concentrations (300 and 30 ng/µL) (A) before, and (B) after colorimetric LAMP reaction. COVID-19 ositive samples (S1, S2, S3. S4. S6, and S8) can be easily discriminated by visual inspection. (C) Distance in color of samples of RNA extracts with respect to negative samples (S5 and S7) in the CIELab space. Distances in color of samples containing 300 ng of nucleic acids (orange bars), or 30 ng of nucleic acids (yellow bars) are presented. (D-E) LAMP amplification products from RNA extracts containing (D) 300 ng/µL and (E) 30 ng/ µL from the same set of COVID (+) patients (S1-S4, S6 and S8), and COVID (-) volunteers (S5 and S7), as revealed by gel electrophoresis experiments. Lanes 1 to 8 contained amplification products from samples S1 to S8. Lane 9 was reserved for the molecular weight ladder. Panel D and E corresponds to portions of the full-length gels presented in supplemental figures S10A and S10B, respectively.

In this reduced set of extracts from nasopharyngeal patient samples, diagnostic results from colorimetric LAMP were completely consistent with RT-qPCR results.

Moreover, discrimination of positive samples even in diluted samples suggests that this colorimetric technique may be useful even in situations where the amount of RNA extracted is low due to improper sampling/extraction or degradation during transportation.

### Concluding remarks

The challenge of point-of-care detection of viral threats is of paramount importance, particularly in underdeveloped regions and in emergency situations (i.e., epidemic outbreaks). In the context of the current COVID-19 pandemic, the availability of testing infrastructure based on RT-qPCR is recognized as a serious challenge around the world. In developing economies (i.e. Latin America, India, and most African countries), the currently available resources for massive COVID-19 testing by RT-qPCR will clearly be insufficient. Even in developed countries, the time to get diagnostic RT-qPCR results from a COVID-19 RT-qPCR test currently ranges from 1 to 5 days.

Clearly, the available PCR labs are overburdened with samples, have too few personnel to conduct the tests, are struggling with backlogs on the instrumentation, and face complicated logistics to transport delicate and infective samples while preserving the cold chain.

Here, we have demonstrated that a simple embodiment of a LAMP reaction, assisted by the use of phenol red as a pH indicator and the use of a simple 3D-printed chamber connected to a water circulator can enable the rapid and highly accurate identification of samples that contain artificial SARS-CoV-2 genetic sequences. We also showed, using synthetic SARS-CoV-2 and a limited number of RNA extracts from patients, that colorimetric LAMP is a quantitative method, comparable to RT-qPCR. Amplification is visually evident, without the need for any additional instrumentation, even at low viral copy numbers. In our experiments with synthetic samples, we observed 100% accuracy in samples containing as few as 625 copies of SARS-CoV-2 genetic material.

Validation of these results using a larger number of real human samples from positive and negative COVID-19 subjects is obviously needed to obtain a full assessment of the potential of this strategy as an alternative to RT-qPCR platforms. However, our results with synthetic samples and with a reduced number of samples containing RNA from human volunteers (8 samples) suggest that this simple strategy may greatly enhance the capabilities for COVID-19 testing in situations where RT-qPCR is not feasible or is unavailable. Recently, other groups have shown that accurate discrimination between COVID-19 positive and negative samples is positive in extraction-free RT-qPCR[52] and colorimetric LAMP implementations[45]. Smyrlaki et al. extensively investigated the performance of an extraction-free RT-qPCR protocol in which saliva or nasopharyngeal samples were heated at 95 °C for 5 minutes for virus inactivation and then directly amplified. Lalli et al. showed successful amplification using a similar colorimetric LAMP protocol in which they heated saliva samples at 50 or 64 °C for 5 minutes optionally adding protease K. These results suggest that extraction-free implementations of amplification methods, including colorimetric LAMP, can successfully identify COVID-19 positive patients from nasopharyngeal and saliva samples. Altogether, this body of evidence suggests that extraction-free colorimetric LAMP provides means for cost-effective massive diagnostics of SARS-CoV-2 and is a promising tool for pandemic contention that deserves further exploration.

## Materials and Methods

### Equipment specifications

We ran several hundred amplification experiments using a colorimetric LAMP method in a 3D-printed incubation chamber designed in house and connected to a conventional water circulator (Figure 1). The design and all dimensional specifications of this chamber have been made available in Supplementary Information (Figures S1, S2; Supplementary File S1). In the experiments reported here, we used a chamber with dimensions of 20 × 5 × 15 cm^3^ and a weight of 0.4 kg (without water). A conventional water circulator (WVR, PA, USA), was used to circulate hot water (set point value at 76 °C) through the 3D-printed chamber for incubation of the Eppendorf PCR tubes (0.2 mL). In this first chamber prototype, twelve amplification reactions can be run in parallel. This concept design is amenable for fabrication in any STL-3D printing platform and may be scaled up to accommodate a larger number of tubes.

We used a blueGel electrophoresis unit, powered by 120 AC volts, to validate the LAMP amplification using gel electrophoresis. Photo-documentation was done using a smartphone camera. We also used a Synergy HT microplate reader (BioTek Instruments, VT, USA) to detect the fluorescence induced by an intercalating reagent in positive samples from the PCR reactions.

### Validation DNA templates

We used plasmids containing the complete N gene from 2019-nCoV, SARS, and MERS as positive controls, with a concentration of 200,000 copies/µL (Integrated DNA Technologies, IA, USA). Samples containing different concentrations of synthetic nucleic acids of SARS-CoV-2 were prepared by successive dilutions from stocks (from 2 × 10^5^ copies to 65 copies). We used a plasmid that contained the gene GP from EBOV as a negative control. The production of this EBOV genetic material has been documented elsewhere by our group [23].

### RNA extracts from human volunteers

In addition, we used 8 samples of RNA extracts from 6 COVID-19 positive and 2 negative subjects, as determined by RT-PCR analysis. Samples were kindly donated by Hospital Alfa, Medical Center, in Guadalupe, Nuevo León, México. Nasopharyngeal samples were collected from two patients after obtaining informed and signed written consent and in complete observance of good clinical practices, the principles stated in the Helsinki Declaration, and applicable lab operating procedures at Hospital Alfa. Every precaution was taken to protect the privacy of sample donors and the confidentiality of their personal information. The experimental protocol was approved on May 20^th^, 2020 by a named institutional committee (Alfa Medical Center, Research Comitte; resolution AMCCI-TECCOVID-001). RNA extraction and purification was conducted at the molecular biology laboratory at Hospital Alfa. The Qiagen QIAamp DSP Viral RNA Mini kit was used for RNA extraction and purification by closely following the directions of the manufacturer.

### Amplification mix

We used WarmStart^®^ Colorimetric LAMP 2× Master Mix (DNA & RNA) from New England Biolabs (MA, USA), and followed the recommended protocol: 12.5 μL Readymix, 1.6 μM FIP primer, 1.6 μM BIP primer, 0.2 μM F3 primer, 0.2 μM B3 primer, 0.4 μM LF primer, 0.4 μM LB primer, 1μL DNA template (∼ 625 to 2 × 10^5^ DNA copies), 1.25 μL EvaGreen^®^ Dye from Biotium (CA, USA), and nuclease-free water to a final volume of reaction 25 μL. This commercial mix contains phenol red as a pH indicator for revealing the shift of pH during LAMP amplification across the threshold of pH=6.8.

### Primers used

Two different sets of LAMP primers, referred to here as α and β, were designed in house using the LAMP primer design software Primer Explorer V5 (http://primerexplorer.jp/lampv5e/index.html). These primers were based on the analysis of alignments of the SARS-Co2 N gene sequences using the software Geneious (Auckland, New Zealand), downloaded from https://www.ncbi.nlm.nih.gov/genbank/sars-cov-2-seqs/#nucleotide-sequences.

Each set, containing six LAMP primers, were used to target two different regions of the sequence of the SARS-Co2 N gene. In addition, for comparison purposes, we conducted PCR amplification experiments using one of the primer sets recommended by the CDC for the standard diagnostics of COVID-19 (i.e., N1 assay) using RT-qPCR. The sequences of our LAMP primers are presented in Table 1. The sequences of the PCR primers (N1) have been reported elsewhere[24,53].

### Amplification protocols

For all LAMP experiments, we performed isothermal heating for 30 or 60 min. In our experiments, we tested three different temperatures: 50, 60, and 65 °C. For PCR experiments, we used a three-sta e protocol consistin o a denaturation sta e at 4 or 5 min, ollo ed y 25 cycles o 4 °C for 20 s, 60 °C for 30 s, and 72 °C for 20 s, and then a final stage at 72 °C for 5 min, for a total duration of 60 min in the miniPCR^®^ thermocycler from Amplyus (MA, USA).

### Documentation of LAMP products

We analyzed 10 μL o each LAMP reaction in a blueGel unit, a portable electrophoresis unit sold by MiniPCR from Amplyus (MA, USA). This is a compact electrophoresis unit (23 × 10 × 7 cm^3^) that weighs 350 g. In these experiments, e analyzed 10 μL o the LAMP product usin 1.2% a arose electrophoresis tris-borate-EDTA buffer (TBE). We used the Quick-Load Purple 2-Log DNA Ladder (NEB, MA, USA) as a molecular weight marker. Gels were dyed with Gel-Green from Biotium (CA, USA) using a 1:10,000 dilution, and a current of 48 V was supplied by the blueGel built-in power supply (AC 100–240V, 50–60Hz).

As an alternative method for detection and reading of the amplification product, we evaluated the amplification products by detecting the fluorescence emitted by a DNA-intercalating agent, the EvaGreen^®^ Dye from Biotium (CA, USA), in a Synergy HT microplate reader (BioTek Instruments, VT, USA). Brie ly, 25 μL o the LAMP reaction was placed in separate wells of a 96-well plate following completion of the LAMP incu ation. A 125 μL volume o nuclease-free water was added to each well for a final sample volume o 150 μL and the samples ere well-mixed by pipetting. These experiments were run in triplicate. The following conditions were used in the microplate reader: excitation of 485/20, emission of 528/20, gain of 75. Fluorescence readings were done from the top at room temperature.

### Color determination by image analysis

We also photographically documented and analyzed the progression of color changes in the positive and negative SARS-CoV-2 synthetic samples during the LAMP reaction time (i.e., from 0 to 50 min). For that purpose, Eppendorf PCR tubes containing LAMP samples were photographed using a smartphone (iPhone, from Apple, USA). We used an application for IOS (Color Companion, freely available at Apple store) to determine the components of color of each LAMP sample in the CIELab color space. Color differences between the positive samples and negative controls were calculated as distances in the CIELab coordinate system according to the following formula:

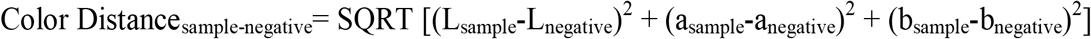

Here L, a, and b are the color components of the sample or the negative control in the CIELab color space (Figure S4).

## Data Availability

All data related to the research described in this manuscript has been made available within the text and figures within the manuscript or as supplemental material.

## Acknowledgments

The authors aknowledge the funding provided by the Federico Baur Endowed Chair in Nanotechnology (0020240I03). EGG acknowledges funding from a doctoral scholarship provided by CONACyT (Consejo Nacional de Ciencia y Tecnología, México). GTdS and MMA acknowledge the institutional funding received from Tecnológico de Monterrey (Grant 002EICIS01). MMA, GTdS, SOMC and IMLM acknowledge funding provided by CONACyT (Consejo Nacional de Ciencia y Tecnología, México) through grants SNI 26048, SNI 256730, SNI 31803, SNI 1056909, respectively. YSZ acknowdges the suppport by the Brigham Research Institute.

## Author participation

E.G. and I.L. conducted most of the the amplification experiments; G.T. and M.A. wrote the manuscript; E.G. I.L. and M.A. prepared the figures; C.G., G.E.G., G.G, and J.A. designed the incubation chamber; A.G. and F.Y. 3D-printed the incubation chamber; I.R. and E.M. conducted the experiments of RNA extraction and purification form human samples at Alfa Medical and INER; Y-S.Z., S.M., G.T., and J.Z. edited the manuscript; J.Z. coordinated the collection of samples and the execution of RT-qPCR experiments at INER, and G.T., S.M., Y-S.Z.; and M.A. designed the study.

## Supporting Information

**Figure S1.** (A) Photograph and (B) rendering of the 3D-printed incubation chamber used in LAMP experiments.

**Figure S2.** Schematic representation of the chamber (different views) showing its relevant dimensions.

**Figure S3.** Commercial plasmid that contains the plasmids containing the complete N gene from 2019-nCoV, SARS, and MERS. We use this plasmid as a SARS-CoV-2 synthetic nucleic acid material in our amplification experiments.

**Figure S4.** *In house* designed plasmid containing the gene that codes for the expression of protein GP from EBOV. This plasmid was added as nucleic acid material in negative controls in our amplification experiments.

**Figure S5.** (A) The colorimetric LAMP method described here was able to identify and amplify synthetic SARS-CoV-2 genetic material in samples containing as few as ∼62 viral copies. (B) Evaluation of the stability and functionality of the LAMP reaction mix at different storage times and temperatures. The reaction mix, which is formulated with LAMP primers and ready for the addition of nucleic acid extracts, is functional and discriminates between positive and negative samples when stored (i) at room temperature for 48 h or (ii) at 4 °C for 72 h.

**Figure S6.** (A) Color analysis conducted on positive and negative SARS-CoV-2 samples contained in Eppendorf PCR tubes (yellow inset) using Color Companion, an app from Apple (downloadable at Apple Store, USA). This app identifies the components of color in a specific location of an image (black circle in the yellow inset) in the CIELab, RGB, HSB, or CMYK spaces. The image can be uploaded using e-mail, airdrop, or Whatsapp. (B) Schematic representation of the CIELab space, a color system where any color can be represented in terms of a point and its coordinates in a 3D space, where L is luminosity, a is the axis between green and red, and b is the axis between yellow and red.

**Figure S7.** (A) The amount of amplification product in LAMP experiments was evaluated by measuring the fluorescence emitted by the amplification product in reactions with an added intercalating agent. Fluorescence readings were conducted in standard 96-well plates using a conventional plate reader. (A) Fluorescence readings, as measured in a commercial plate reader, for different dilutions of SARS-CoV-2 synthetic DNA templates. Results using two different LAMP primer sets are shown: set α (indicated in blue), and set β (indicated in red).

**Figure S8.** Images of the full-length gels from which Figure 3B and 3C were obtained. (A, B) Agarose gel electrophoresis of DNA amplification products generated by targeting two different regions of the sequence coding for SARS-Co2 N protein. Two different primer sets were used: (B) primer set α, and (C) β. The initial template amount was gradually decreased from left to right: 2.0 × 105 DNA copies (lane 1), 4.0 × 104 copies, (lane 2), 1.0 × 104 copies (lane 3), 2.5 × 103 copies (lane 4), 625 copies (lane 5), negative control (lane 6), and molecular weight ladder (lane 7).

**Figure S9.** Image of the full-length gel from which Figure 6B was obtained. LAMP amplification products from RNA extracts (lane 1 and 2) and cDNA (lane 3 and 4) from a COVID (+) patient, and a COVID (-) volunteer (lane 5), as revealed by gel electrophoresis experiments. A molecular weight ladder is shown in lane 6.

**Figure S10.** Image of the full-length gels from which Figure 7D and 7E were obtained. (A-B) LAMP amplification products from RNA extracts containing (D) 300 ng/µL and (E) 30 ng/ µL from the same set of COVID (+) patients (S1-S4, S6 and S8), and COVID (-) volunteers (S5 and S7), as revealed by gel electrophoresis experiments. Lanes 1 to 8 contained amplification products from samples S1 to S8. Lane 9 was reserved for the molecular weight ladder.

